# Long-term mortality outcomes among immunotherapy recipients treated with dupilumab for the management of cutaneous immune-related adverse events

**DOI:** 10.1101/2025.01.07.25320156

**Authors:** Sara Khattab, Guihong Wan, Suzanne Xu, Cameron Moseley, Matthew Tran, Emma Beagles, Chuck Lin, Bonnie Leung, Marjan Azin, Ninghui Hao, Kerry L. Reynolds, Shadmehr Demehri, Nicole R. LeBoeuf, Yevgeniy R. Semenov

## Abstract

**Background:** Dupilumab has been added to National Cancer Comprehensive Network (NCCN) guidelines as a therapeutic strategy for managing certain cutaneous immune-related adverse events (cirAEs) from immune checkpoint inhibitor (ICI) therapy. However, little is known about the implications of dupilumab for cancer outcomes in this population. In this multi-institutional study, we evaluate the impact of dupilumab treatment on survival among ICI recipients.

**Methods:** We conducted a muti-institutional retrospective cohort study of ICI recipients from the Mass General Brigham Healthcare System and Dana-Farber Cancer Institute. The dupilumab group was compared to two control groups who did not receive dupilumab: with and without cirAEs (control groups 1 and 2, respectively) that were 1:2 matched on sex, race, age at ICI initiation, Charlson Comorbidity Score, year of ICI initiation, and ICI type. Manual chart review was performed to obtain cirAE characteristics, systemic glucocorticoid use, dupilumab treatment, vital status, and last contact date. Time-varying multivariable Cox proportional hazards regressions were used to evaluate the impact of dupilumab on overall survival, adjusted for sex, race, age at ICI initiation, ICI type, Charlson Comorbidity Index score, cancer type, cancer stage at ICI initiation, and systemic glucocorticoid use.

**Results:** A total of 53 cirAE patients treated with dupilumab were compared to two control groups of 106 patients each. Most patients receiving dupilumab demonstrated either complete or partial resolution of their cirAE (88.7%). In multivariable modeling, the overall survival of the dupilumab group was not significantly different from control group 1 (HR=0.74, 95% CI:0.35-1.60, p=0.5) or control group 2 (HR=0.70, 95% CI:0.32-1.51, p=0.4). However, the use of systemic glucocorticoids within two years after ICI initiation was associated with poorer overall survival when comparing the dupilumab group to control group 1 (HR=2.03, 95% CI:1.04-3.96, p=0.039) and control group 2 (HR=2.21, 95% CI:1.25-3.91, p=0.006).

**Conclusions:** This study suggests that dupilumab is an effective therapeutic option for recalcitrant cirAEs and does not adversely impact mortality. Due to the observed detrimental effects of systemic glucocorticoid therapy, this study supports the need to shift away from systemic glucocorticoid immunosuppression and towards targeted immune modulators for irAE management that are less detrimental to ICI response.

**• What is already known on this topic:** Current guidelines recommend the use of dupilumab in the treatment of certain moderate to severe cutaneous immune related adverse events (cirAE) and systemic glucocorticoids for others. Previous studies have shown dupilumab to be effective for steroid refractory cirAEs;^1^ however, the impact dupilumab on survival outcomes among recipients of immune checkpoint inhibitor therapy (ICI) remains under studied.

**• What this study adds:** This study concludes that dupilumab is an effective modality to treat cirAEs, with 88.7% of patients responding to treatment. Additionally, this study demonstrates a 206-day average delay from cirAE onset to dupilumab treatment suggesting the need for more timely consideration of this therapeutic option. Finally, our results demonstrated that dupilumab does not increase mortality among ICI recipients.

**• How this study might affect research, practice or policy:** The results of this study suggest that use of dupilumab in the treatment of cirAEs is effective and does not adversely impact mortality in the cancer population. Based on these findings, clinicians should consider dupilumab treatment for cirAEs in the appropriate clinical setting. Moreover, this study provides further evidence for the use of targeted immune modulators as preferred over more commonly utilized broad-based glucocorticoid immunosuppression for the management of immune related adverse events in the setting of ICI therapy.

## Introduction

Immune checkpoint inhibitor (ICI) therapy has revolutionized cancer treatment but is associated with morbid and potentially life-threatening toxicities known as immune-related adverse events (irAEs). Of these, cutaneous immune related adverse events (cirAEs) are the most common, occurring in up to 40% of ICI recipients. Current National Cancer Comprehensive Network (NCCN) version 1.2024 guidelines for the management of cirAEs of different morphologies range from the use of topical steroids and oral antihistamines for low grade eruptions to holding immunotherapy and initiating high dose systemic immunosuppressive agents, typically systemic glucocorticoid therapy, for high grade eruptions.^2^ Though there is not yet consensus regarding the use of systemic immunosuppression in patients receiving ICI treatment for cancer care,^3–5^ there is a growing body of evidence suggesting that the use of systemic glucocorticoids in ICI-treated patients is detrimental to survival outcomes,^6,7^ especially early in the course of ICI therapy.^8^ In response to this concern, increasing attention has been given to considering more targeted immune-modulating approaches for the management of irAEs, which are hypothesized to be less likely to blunt the anti-tumor effect of ICIs than broader systemic glucocorticoid immunosuppression.

Evidence of this approach notes the recent inclusion of dupilumab, currently approved by the United States Food and Drug Administration for the management of atopic dermatitis and prurigo nodularis, in the NCCN guidelines for the management of certain cirAEs. Dupilumab is a monoclonal antibody that inhibits interleukin 4 (IL-4) and interleukin 13 (IL-13) cytokine signaling and prevents the release of downstream immunoglobulin E (IgE), which plays an important role in immune mediated allergic processes, primarily in the type 2 inflammatory pathway.^9,10^ Though these guidelines propose the use of dupilumab for the management of moderate to severe bullous eruptions and severe pruritus in the setting of ICI therapy,^2^ it has been rapidly adopted as an off-label therapeutic strategy across a wide range of cirAEs. However, though several recent studies have demonstrated the efficacy of dupilumab in the management of several specific morphologies of cirAEs, these have been limited to case reports^11–14^ and single institutional cohorts^1,15,16^ without inclusion of comparator populations and long-term follow-up to evaluate the specific impact of dupilumab on ICI outcomes. As a result, there is no available data on the long-term impact of dupilumab on mortality among ICI recipients.

In this multi-institutional retrospective cohort study, our primary aim is to evaluate overall survival outcomes among ICI recipients treated with dupilumab. Our secondary aim includes evaluating the efficacy of dupilumab in the management of cirAEs. To our knowledge, this is the largest cohort of ICI recipients treated with dupilumab to date and the first to explicitly evaluate survival outcomes by comparison to robust non-dupilumab treated comparator populations of ICI recipients.

## Methods

We conducted a muti-institutional retrospective cohort study of ICI recipients who received dupilumab therapy for the management of cirAEs between September 27, 2017, and December 08, 2023, at the Mass General Brigham Healthcare System and the Dana-Farber Cancer Institute (MGBD). Figure 1 presents the population included in this study. We extracted patient demographic and medical history information from the MGBD Research Patient Data Registry^17^ and the Enterprise Data Warehouse^18^ using the same approaches as in our recently published studies^19–21^ and in alignment with the published guidelines on defining cirAEs^22^.

**Figure 1.**
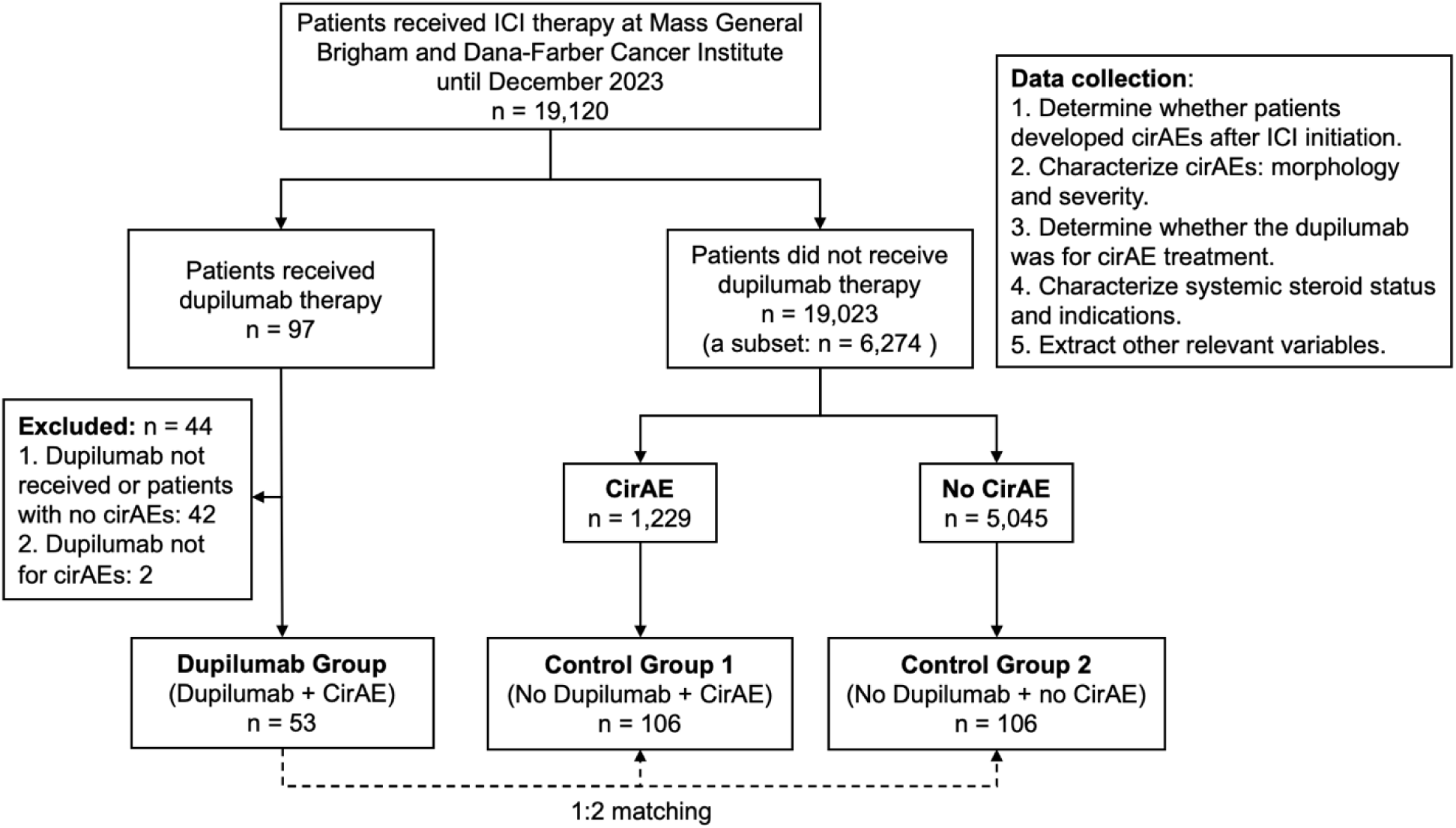
The study population and data collection This study identified all ICI recipients who developed cirAEs and received Dupilumab for managing cirAEs as the case group (the dupilumab group). To demonstrate the robustness of this study, the dupilumab group was compared to two control groups that were identified using 1:2 matching based on sex, race, age at ICI initiation, Charlson Comorbidity Score, year of ICI start, and ICI type. The first control group included 106 ICI recipients who developed cirAEs and were not treated with Dupilumab; the second control group included 106 ICI recipients who did not experience cirAEs and were not treated with Dupilumab. ICI: Immune Checkpoint Inhibitor; CirAE: Cutaneous immune-related adverse event.

Manual chart review was conducted to extract cirAE characteristics (cirAE status – yes or no, cirAE onset date, cirAE morphology, and cirAE severity), dupilumab variables (dupilumab status – yes or no, dupilumab indication, dupilumab start and end date, treatments before dupilumab initiation, and dupilumab response), immunosuppression variables (systemic glucocorticoid use – yes or no, start date, and indication within 24 months after ICI initiation), absolute eosinophil count before and after dupilumab start, and patient outcomes (vital status, and date of last contact). Manual chart review to identify cirAEs was conducted in accordance with our previously published approaches.^19–21^ Briefly, a likelihood score between one to four was assigned to each cutaneous eruption in the setting of ICI use, with 1 representing that the eruption is highly unlikely to be secondary to ICI treatment and 4 representing that the eruption is highly likely to be secondary to ICI treatment. Cutaneous eruptions with a likelihood score of 3 or 4 were considered as cirAEs in this study.

High dose systemic glucocorticoid use was defined as treatment with a glucocorticoid of 10mg prednisone equivalent daily for at least 7 consecutive days, which is consistent with prior literature.^23^ CirAE severity was graded using Common Terminology Criteria for Adverse Events (CTCAE) version 5.0.^24^ CirAE morphology was documented based on clinical and histologic confirmation, whenever available. If histologic confirmation was not available, the morphology was documented based on clinical assessment by the dermatology team. Response to dupilumab was measured by comparing the CTCAE grade of the cirAE before and after the use of dupilumab. Patients whose cirAE became grade 0 were considered complete responders. Patients whose cirAE became grade 1 were considered partial responders. Both complete responders and partial responders were categorized as responders.

Patients who did not ultimately receive dupilumab treatment or who were given dupilumab for treatment of a condition besides a cirAE were excluded from the study population. The retained ICI recipients who were treated with dupilumab for management of cirAEs (dupilumab group) were then compared to two control groups that were identified by 1:2 best matching based on sex, race, age at ICI initiation, Charlson Comorbidity Score (CCS), year of ICI start, and ICI type using the "matchControls" function in the R package e1071 version 1.7-14. The first control group included ICI recipients who developed cirAEs but were not treated with dupilumab (control group 1). The second group included ICI recipients who did not experience cirAEs and were not treated with dupilumab (control group 2). Because the development of cirAEs has been associated with improved survival in the setting of ICI therapy,^19,23,25^ we controlled for the presence of cirAEs by matching with patients who developed cirAEs but did not use dupilumab (control group 1). The second control group was used to examine the impact of dupilumab on overall survival independent of the presence of cirAEs.

We used Pearson’s chi-squared test for categorical variables and t-test for continuous variables to compare groups. We used an alpha of 0.05 as the significance threshold. To account for immortal time bias,^26^ we performed time-varying Cox proportional hazards modeling, adjusting for sex, race, age at ICI initiation, ICI type, CCS, cancer type, cancer stage at ICI initiation, and systemic glucocorticoid use within two years after ICI initiation. Both dupilumab and high dose systemic glucocorticoid use were considered as time-varying covariates. The proportional hazards assumption of was examined using the "cox.zph" function in the R package survival version 3.5-7. All statistical analyses were conducted in R version 4.3.2.

## Results

A total of 53 ICI recipients who received dupilumab for the management of cirAEs were included and were matched to 106 ICI recipients with cirAEs but no dupilumab treatment and 106 ICI recipients without cirAEs and without dupilumab therapy (Figure 1). The characteristics of the dupilumab group and the two control groups are presented in Table 1. Comparing the dupilumab group and the control group 1, there were no significant differences in mortality status, follow-up duration, sex, race, age at ICI initiation, year of ICI initiation, ICI type, CCS, cancer stage at ICI initiation, and high dose systemic glucocorticoid use within two years after initiation of ICI therapy (p>0.05). Comparing the dupilumab group and the control group 2, there were significant differences in mortality status (18.9% vs 55.7%, p<0.001), follow-up duration (median: 961 vs 416 days, p<0.001), CCS (median: 1 vs 2, p=0.023), and high dose systemic glucocorticoid use within two years after initiation of ICI therapy (71.7% vs 48.1%, p=0.008).

**Table 1.** Characteristics of the study population.

|  | **Dupilumab**<br>**(N=53)** | **Control 1**<br>**(N=106)** | **Control 2**<br>**(N=106)** | **P-value 1 <sup>a</sup>** | **P-value 2 <sup>a</sup>** |
| --- | --- | --- | --- | --- | --- |
| **Mortality status** |  |  |  |  |  |
| Alive | 43 (81.1%) | 69 (65.1%) | 47 (44.3%) | 0.057 | <0.001 |
| Dead | 10 (18.9%) | 37 (34.9%) | 59 (55.7%) |  |  |
| **Follow-up duration, days** |  |  |  |  |  |
| Median [Q1, Q3] | 961 [531, 1240] | 718 [432, 1090] | 416 [185, 791] | 0.114 | <0.001 |
| **Sex** |  |  |  |  |  |
| Female | 18 (34.0%) | 35 (33.0%) | 35 (33.0%) | >0.9 | >0.9 |
| Male | 35 (66.0%) | 71 (67.0%) | 71 (67.0%) |  |  |
| **Race** |  |  |  |  |  |
| White | 47 (88.7%) | 99 (93.4%) | 101 (95.3%) | 0.474 | 0.224 |
| Other/Unavailable | 6 (11.3%) | 7 (6.6%) | 5 (4.7%) |  |  |
| **Age at ICI initiation, years** |  |  |  |  |  |
| Median [Q1, Q3] | 67 [61, 75] | 66 [61, 74] | 67.5 [62, 74] | 0.360 | 0.848 |
| **Year of ICI initiation** |  |  |  |  |  |
| <2018 | 2 (3.8%) | 5 (4.7%) | 5 (4.7%) | >0.9 | >0.9 |
| 2018 | 5 (9.4%) | 12 (11.3%) | 10 (9.4%) |  |  |
| 2019 | 5 (9.4%) | 9 (8.5%) | 9 (8.5%) |  |  |
| 2020 | 14 (26.4%) | 28 (26.4%) | 28 (26.4%) |  |  |
| 2021 | 12 (22.6%) | 21 (19.8%) | 24 (22.6%) |  |  |
| 2022 | 12 (22.6%) | 25 (23.6%) | 24 (22.6%) |  |  |
| 2023 | 3 (5.7%) | 6 (5.7%) | 6 (5.7%) |  |  |
| **ICI type <sup>b</sup>** |  |  |  |  |  |
| Combination | 18 (34.0%) | 38 (35.8%) | 29 (27.4%) | >0.9 | 0.499 |
| PD-1/PD-L1 | 35 (66.0%) | 68 (64.2%) | 77 (72.6%) |  |  |
| **CCS** |  |  |  |  |  |
| Median [Q1, Q3] | 1 [0, 3] | 2 [0, 3] | 2 [1, 3] | 0.197 | 0.023 |
| **Cancer stage at ICI <sup>c</sup>** |  |  |  |  |  |
| IV | 37 (69.8%) | 74 (69.8%) | 85 (80.2%) | 0.292 | 0.341 |
| III | 11 (20.8%) | 28 (26.4%) | 14 (13.2%) |  |  |
| Other | 5 (9.4%) | 4 (3.8%) | 7 (6.6%) |  |  |
| **Cancer type** |  |  |  |  |  |
| Melanoma | 13 (24.5%) | 50 (47.2%) | 30 (28.3%) | 0.013 | 0.887 |
| Genitourinary | 16 (30.2%) | 28 (26.4%) | 29 (27.4%) |  |  |
| Head and neck | 8 (15.1%) | 8 (7.5%) | 13 (12.3%) |  |  |
| Thoracic | 8 (15.1%) | 16 (15.1%) | 21 (19.8%) |  |  |
| Other | 8 (15.1%) | 4 (3.8%) | 13 (12.3%) |  |  |
| **Systemic glucocorticoids use within two years of ICI start <sup>d</sup>** |  |  |  |  |  |
| Yes | 38 (71.7%) | 64 (60.4%) | 51 (48.1%) | 0.220 | 0.008 |
| No | 15 (28.3%) | 42 (39.6%) | 55 (51.9%) |  |  |
| **Systemic glucocorticoid reason <sup>d</sup>** |  |  |  |  |  |
| CirAEs | 26 (49.1%) | 12 (11.3%) | 0 (0%) | <0.001 | <0.001 |
| Other irAEs | 10 (18.9%) | 42 (39.6%) | 28 (26.4%) |  |  |
| Cancer Palliation | 2 (3.8%) | 7 (6.6%) | 12 (11.3%) |  |  |
| Other | 0 (0%) | 3 (2.8%) | 11 (10.4%) |  |  |
| **ICI to systemic glucocorticoid <sup>d</sup>, days** |  |  |  |  |  |
| Median [Q1, Q3] | 253 [74, 518] | 140 [67.8, 267] | 168 [47.5, 263] | 0.046 | 0.010 |

<sup>a</sup> P-value 1: comparison between the dupilumab group and the control 1 group. P-value 2: comparison between the dupilumab group and the control 2 group. <sup>b</sup> Combination therapy of CTLA-4 and PD-1/PD-L1. <sup>c</sup> Other includes the cases where the corresponding cancers were not staged based on AJCC criteria. <sup>d</sup> For these variables, we consider the systemic glucocorticoid use within two years after the initiation of ICI therapy.
ICI: Immune checkpoint inhibitor; irAEs: immune-related adverse events; cirAEs: cutaneous immune-related adverse events; CCS: Charlson comorbidity score; CTLA-4: Cytotoxic T-Lymphocyte Associated Protein 4; PD-1: Programmed Death-1; PD-L1: Programmed Death-Ligand 1; Q1: the first quartile; Q3: the third quartile.

Among the 38 patients who received high dose systemic glucocorticoids within two years after initiation of ICI therapy in the dupilumab group, 81.6% (31 patients) received systemic glucocorticoids before dupilumab treatment to manage either cutaneous irAEs (24 patients) or non-cutaneous irAEs (7 patients). The remaining 18.4% (7 patients) who were treated with high dose systemic glucocorticoids, received it after initiating dupilumab therapy, for the management of either cutaneous irAEs (3 patients), non-cutaneous irAEs (1 patient), cerebral edema from brain metastases (2 patients), or worsening pre-existing cough (1 patient).

Among the dupilumab group, 30.2% (16/53) of patients developed an initial cirAE presentation before a subsequent cirAE for which treatment with dupilumab was indicated. Among these 16 patients, dupilumab-treated cirAEs had more morphologic specificity (8 patients: from unspecific rash to eczematous dermatitis, lichenoid dermatitis, or bullous pemphigoid; 4 patients: from pruritus to eczematous dermatitis or lichenoid dermatitis; 3 patients: from maculopapular eruption to bullous pemphigoid or sclerodermoid reaction with morphea-profunda; and 1 patient: lichenoid dermatitis to lichenoid dermatitis and bullous pemphigoid) and the grade was higher (greater than grade 1: 98.1% vs 75.5%, p=0.003) by comparison to the first cirAE presentation. Table 2 presents the cirAE severity and morphologies. Moreover, compared to control group 1, the first cirAE presentation for the dupilumab group was more severe (greater than grade 1: 75.5% vs 50.9%, p=0.012). Among all 53 dupilumab-treated patients, 22 (41.5%) had eczematous dermatitis, 14 (26.4%) had bullous pemphigoid, 7 (13.2%) had lichenoid dermatitis, 5 (9.4%) had morbilliform drug eruptions, 3 (5.7%) had mixed morphology (which consisted of lichenoid dermatitis/morbilliform drug eruption, lichenoid dermatitis/bullous pemphigoid, and lichenoid dermatitis/eczematous dermatitis), and 2 (3.8%) had other morphologies (radiation induced ICI exacerbated morphea and sclerodermoid reaction with morphea profunda) (Table 2). The median time from ICI start to first cirAE onset was 63 vs 54.5 days (p=0.395) for dupilumab and control group 1 cohorts, respectively. The median duration from ICI start to the onset of dupilumab-treated cirAEs and to the initiation of dupilumab treatment was 146 and 352 days, respectively. The median duration of dupilumab treatment was 230 days.

**Table 2.** Severity and morphology of cirAEs and eosinophil count.

|  | **Dupilumab**<br>**(N=53)** | **Control 1**<br>**(N=106)** | **P-value** |
| --- | --- | --- | --- |
| **Severity of the first cirAE** |  |  |  |
| 1 | 13 (24.5%) | 52 (49.1%) | 0.012 |
| 2 | 32 (60.4%) | 43 (40.6%) |  |
| 3 | 8 (15.1%) | 11 (10.4%) |  |
| **Severity of the dupilumab-treated cirAE** |  |  |  |
| 1 | 1 (1.9%) | N/A |  |
| 2 | 42 (79.2%) | N/A |  |
| 3 | 10 (18.9%) | N/A |  |
| **Morphology of the first cirAE** |  |  |  |
| Eczematous dermatitis | 15 (28.3%) | 7 (6.6%) | <0.001 |
| Bullous pemphigoid | 10 (18.9%) | 3 (2.8%) |  |
| Morbilloform drug eruption | 5 (9.4%) | 7 (6.6%) |  |
| Lichenoid dermatitis | 5 (9.4%) | 7 (6.6%) |  |
| Lichenoid dermatitis, eczematous dermatitis | 1 (1.9%) | 0 (0%) |  |
| Lichenoid dermatitis, morbilliform drug eruption | 1 (1.9%) | 0 (0%) |  |
| Radiation induced morphea ICI exacerbated | 1 (1.9%) | 0 (0%) |  |
| Maculopapular eruption | 3 (5.7%) | 20 (18.9%) |  |
| Pruritus | 4 (7.5%) | 16 (15.1%) |  |
| Psoriasiform eruption | 0 (0%) | 6 (5.7%) |  |
| Vitiligo | 0 (0%) | 7 (6.6%) |  |
| Rash, NOS | 8 (15.1%) | 33 (31.1%) |  |
| **Morphology of the dupilumab-treated cirAE** |  |  |  |
| Eczematous dermatitis | 22 (41.5%) | N/A |  |
| Bullous pemphigoid | 14 (26.4%) | N/A |  |
| Lichenoid dermatitis | 7 (13.2%) | N/A |  |
| Morbilloform drug eruption | 5 (9.4%) | N/A |  |
| Lichenoid dermatitis, bullous pemphigoid | 1 (1.9%) | N/A |  |
| Lichenoid dermatitis, eczematous dermatitis | 1 (1.9%) | N/A |  |
| Lichenoid dermatitis, morbilliform drug eruption | 1 (1.9%) | N/A |  |
| Radiation induced morphea ICI exacerbated | 1 (1.9%) | N/A |  |
| Sclerodermoid reaction with morphea-profunda | 1 (1.9%) | N/A |  |
| **ICI to the first cirAE, days** |  |  |  |
| Median [Q1, Q3] | 63 [15, 198] | 54.5 [21, 155] | 0.395 |
| **ICI to the dupilumab-treated cirAE, days** |  |  |  |
| Median [Q1, Q3] | 146 [21, 414] | N/A |  |
| **ICI to the dupilumab initiation, days** |  |  |  |
| Median [Q1, Q3] | 352 [184, 585] | N/A |  |
| **Dupilumab duration, days** |  |  |  |
| Median [Q1, Q3] | 230 [124, 418] | N/A |  |
| **Absolute eosinophil count before dupilumab** |  |  |  |
| Median [Q1, Q3] | 0.39 [0.18, 0.76] | N/A |  |
| **Absolute eosinophil count after dupilumab** |  |  |  |
| Median [Q1, Q3] | 0.16 [0.07, 0.26] | N/A |  |
| **CBC before dupilumab, days** |  |  |  |
| Median [Q1, Q3] | 26 [18.5, 61] | N/A |  |
| **CBC after dupilumab, days** |  |  |  |
| Median [Q1, Q3] | 140 [72.5, 219] | N/A |  |

CirAE: Cutaneous immune-related adverse event; NOS: Not otherwise specified; CBC: Complete blood count; Q1: the first quartile; Q3: the third quartile.

Of the 53 patients treated with dupilumab, 33 (62.3%) were complete responders, 14 (26.4%) were partial responders, and 6 (11.3%) were non-responders (Table S2). Among single morphologies of cirAEs that were treated with dupilumab, complete response was highest for morbilliform drug eruptions (80%), followed by eczematous eruptions (63.6%). Non-response was highest for bullous pemphigoid (21.4%). All patients with lichenoid eruptions and other eruptions in this cohort had either a complete response or partial response to dupilumab.

Therefore, the response rate to dupilumab treatment was 88.7% (47/53). There was a significant decrease in absolute eosinophil count before and after dupilumab treatment (median: 0.39 vs 0.16, p<0.001) (Table 2).

All patients who received dupilumab had failed prior first-line therapy for the management of their cirAE. Table 3 presents the details of treatments patients received before starting dupilumab for managing cirAEs. In the dupilumab group, 100% (53 patients) received topical treatments, 69.8% (37 patients) were treated with antihistamines, and 66.0% (35 patients) received high-dose (24 patients) or low-dose (11 patients) systemic glucocorticoids for managing cirAEs before the initiation of dupilumab therapy. In the multivariable time-varying Cox proportional hazards models (Table 4), the overall survival of the dupilumab group was not significantly different from control group 1 (HR=0.74, 95% CI:0.35-1.60, p=0.5) or control group 2 (HR=0.70, 95% CI:0.32-1.51, p=0.4). The use of high-dose systemic glucocorticoids within two years following ICI initiation was associated with poorer overall survival (HR=2.03, 95% CI:1.04-3.96, p=0.039) in the regression comparing the dupilumab group to the control group 1 and (HR=2.21, 95% CI:1.25-3.91, p=0.006) in the regression comparing the dupilumab group to the control group 2. Cox modeling assumptions held globally and separately for each covariate in the two models (p>0.05).

**Table 3.** Treatments for cirAEs before dupilumab.

| Treatments for cirAEs before dupilumab <sup>a</sup> | The dupilumab group<br>(N = 53) |  |
| --- | --- | --- |
| Topical treatment alone | 4 (7.5%) | 53<br>(100%) |
| Topical and other treatments | 49 (92.5%) |  |
| Antihistamines | 4 (7.5%) | 12<br>(22.6%) |
| Antihistamines, Anticonvulsants | 2 (3.8%) |  |
| Antihistamines, Anticonvulsants, Phototherapy | 1 (1.9%) |  |
| Antihistamines, IVIg | 1 (1.9%) |  |
| Antihistamines, IVIg, Oral antibiotics, Biologics | 1 (1.9%) |  |
| Antihistamines, Phototherapy | 2 (3.8%) |  |
| Antihistamines, Phototherapy, Oral retinoid | 1 (1.9%) |  |
| Antihistamines, High-dose glucocorticoids | 6 (11.3%) | 25<br>(47.2%) |
| Antihistamines, High-dose glucocorticoids, Anticonvulsants | 5 (9.4%) |  |
| Antihistamines, High-dose glucocorticoids, Anticonvulsants, Oral systemic | 1 (1.9%) |  |
| Antihistamines, High-dose glucocorticoids, Anticonvulsants, Phototherapy | 2 (3.8%) |  |
| Antihistamines, High-dose glucocorticoids, Biologics | 1 (1.9%) |  |
| Antihistamines, High-dose glucocorticoids, Oral systemic, IVIg | 1 (1.9%) |  |
| Antihistamines, High-dose glucocorticoids, Oral systemic | 1 (1.9%) |  |
| Antihistamines, High-dose glucocorticoids, Phototherapy, Oral retinoid | 1 (1.9%) |  |
| Antihistamines, Low-dose glucocorticoids, Anticonvulsants, Oral antibiotics, Oral vitamin | 1 (1.9%) |  |
| Antihistamines, Low-dose glucocorticoids, Anticonvulsants, Phototherapy | 1 (1.9%) |  |
| Antihistamines, Low-dose glucocorticoids | 2 (3.8%) |  |
| Antihistamines, Low-dose glucocorticoids, Oral retinoid | 2 (3.8%) |  |
| Antihistamines, Low-dose glucocorticoids, Phototherapy, Opioid antagonist | 1 (1.9%) |  |
| High-dose glucocorticoids | 2 (3.8%) | 10<br>(18.9%) |
| High-dose glucocorticoids, Anticonvulsants | 1 (1.9%) |  |
| High-dose glucocorticoids, Anticonvulsants, Oral systemic, Hemorrhagic agents | 1 (1.9%) |  |
| High-dose glucocorticoids, Oral antibiotics | 1 (1.9%) |  |
| High-dose glucocorticoids, Oral retinoid | 1 (1.9%) |  |
| Low-dose glucocorticoids | 3 (5.7%) |  |
| Low-dose glucocorticoids, Oral retinoid | 1 (1.9%) |  |
| Anticonvulsants | 1 (1.9%) | 1 (1.9%) |
| Phototherapy, Oral retinoid | 1 (1.9%) | 1 (1.9%) |

<sup>a</sup> All 53 patients in the dupilumab group received topical treatments that were not presented in all rows.

**Table 4.** Time-varying Cox proportional hazards models for overall survival.

|  | **Comparison 1 <sup>a</sup>** |  |  |  | **Comparison 2 <sup>b</sup>** |  |  |
| --- | --- | --- | --- | --- | --- | --- | --- |
|  | **HR** | **95% CI** | **p-value** |  | **HR** | **95% CI** | **p-value** |
| **Dupilumab** | 0.74 | 0.35, 1.60 | 0.5 |  | 0.70 | 0.32, 1.51 | 0.4 |
| **Systemic glucocorticoid** | 2.03 | 1.04, 3.96 | 0.039 |  | 2.21 | 1.25, 3.91 | 0.006 |
| **Age at ICI initiation** | 1.03 | 0.99, 1.06 | 0.11 |  | 1.01 | 0.98, 1.03 | 0.6 |
| **CCS** | 1.10 | 0.96, 1.27 | 0.2 |  | 1.18 | 1.06, 1.32 | 0.003 |
| **Cancer stage at ICI <sup>c</sup>** |  |  |  |  |  |  |  |
| IV | — | — |  |  | — | — |  |
| III and other | 0.75 | 0.37, 1.54 | 0.4 |  | 0.74 | 0.38, 1.42 | 0.4 |
| **Cancer type** |  |  |  |  |  |  |  |
| Melanoma | — | — |  |  | — | — |  |
| Genitourinary | 2.13 | 0.95, 4.77 | 0.066 |  | 1.32 | 0.57, 3.07 | 0.5 |
| Head and neck | 1.98 | 0.69, 5.65 | 0.2 |  | 3.11 | 1.29, 7.47 | 0.011 |
| Thoracic | 1.49 | 0.55, 3.98 | 0.4 |  | 2.04 | 0.81, 5.13 | 0.13 |
| Other | 2.67 | 0.77, 9.22 | 0.12 |  | 3.01 | 1.20, 7.56 | 0.019 |
| **ICI type** |  |  |  |  |  |  |  |
| Combination | — | — |  |  | — | — |  |
| PD-1/PD-L1 | 0.89 | 0.44, 1.81 | 0.8 |  | 0.69 | 0.39, 1.22 | 0.2 |
| **Race** |  |  |  |  |  |  |  |
| White | — | — |  |  | — | — |  |
| Other | 1.66 | 0.65, 4.24 | 0.3 |  | 1.56 | 0.62, 3.92 | 0.3 |
| **Sex** |  |  |  |  |  |  |  |
| Female | — | — |  |  | — | — |  |
| Male | 0.66 | 0.35, 1.22 | 0.2 |  | 1.24 | 0.69, 2.23 | 0.5 |

<sup>a</sup> Comparison 1: comparison between the dupilumab group and the control 1 group.
<sup>b</sup> Comparison 2: comparison between the dupilumab group and the control 2 group.
<sup>c</sup> Other includes the cases where the corresponding cancers were not staged based on AJCC criteria. Due to sample size, we combined them with patients with stage III cancers at ICI initiation in the models.
HR: Hazard Ratio; CI: Confidence Interval; CCS: Charlson Comorbidity Score; PD-1: Programmed Death-1; PD-L1: Programmed Death-Ligand 1

## Discussion

This multi-institutional retrospective cohort study suggests that dupilumab is an effective treatment modality for recalcitrant cirAEs of various morphologies and that its use does not adversely impact mortality among ICI recipients. Notably, the survival trend associated with dupilumab was protective of mortality but did not reach statistical significance in this study due to insufficient sample size of the dupilumab population to demonstrate this protective effect.

Interestingly, a recent study has suggested that dupilumab may enhance response to ICI treatment in ICI-resistant cancers; in six patients with non-small cell lung cancer with progressive disease while on PD-1 or PDL-1 receptor inhibiting immunotherapy, patients were given adjunct dupilumab in addition to their continued ICI regimens.^27^ One of the six patients experienced near complete response following addition of dupilumab.^27^ Similarly, though not reaching statistical significance, our results suggest that the use of dupilumab for cirAE management may be associated with a protective mechanism in the ICI population. Additional studies with larger cohorts are necessary to further elucidate this potential relationship. Our study demonstrates that dupilumab does not increase the risk of mortality in this population and adds valuable data to aid oncologists and dermatologists in guiding their therapeutic selection and counseling patients about the long-term implications of dupilumab use in the setting of ICI therapy. Additionally, this is the largest study of dupilumab efficacy in the ICI-treated population and the first study to explicitly explore its long-term safety profile in an ICI-treated population.

The utility of dupilumab in the management of cirAEs has previously been reported, and our conclusions confirm these findings.^1^ Our results demonstrated an 88.7% response rate to dupilumab therapy among ICI recipients across a broad range of cirAE morphologies, demonstrating that this therapeutic strategy is highly effective. Prior studies have reported an 87% cirAE response rate to dupilumab use,^1^ which our findings independently validate in a larger multi-institutional cohort. We also further stratified responders by cirAE morphology and found that although dupilumab is effective for the management of multiple different morphologies of cirAEs, its efficacy varies across morphologies. For instance, among individual morphologies of cirAEs, dupilumab demonstrated the highest non-response for the management of bullous eruptions and the highest complete response rate for the management of morbilliform drug eruptions followed by eczematous eruptions. Interestingly, all patients with lichenoid and other eruptions achieved partial or complete response with a 90% rate of complete response in patients with mixed morphologies. As a result, this study suggests that the use of dupilumab in the treatment of lichenoid, other, and mixed morphologies could also be useful and that clinicians should consider broadening the indications for which they use dupilumab, despite the current absence of these morphologies in the NCCN guidelines for the management of irAEs.^2^

Additionally, these results confirm that the use of high dose systemic glucocorticoid immunosuppression within two years of immunotherapy initiation is associated with detrimental effects on overall survival, which previous studies have also reported.^6,28^ Our findings also indicate a significant delay from the time of dupilumab treated cirAE start to dupilumab initiation of 206 days. We suspect that this delay may be due to several issues including time to dermatology referral and insurance approval of dupilumab. Based on these findings, we encourage early referral to dermatology for patients experiencing cirAEs.

Our study suggests that dupilumab can be used in the management of treatment-refractory cirAEs without impacting survival. Clinicians should consider using dupilumab in the management cirAEs not responsive to topical therapies as a safer alternative to the more commonly utilized systemic glucocorticoid immunosuppression and should counsel patients that this therapeutic strategy does not adversely impact their ICI outcomes. Additionally, this study demonstrates favorable response to dupilumab treatment across several cirAE morphologies, including non-specific morphologies, and clinicians should discuss this treatment option with their cirAE patients with difficult to classify rashes. This study provides further support for the need to shift the paradigm of irAE management from reliance on systemic glucocorticoid immunosuppression, which may dampen the desired immune response in the setting of ICI treatment, toward more targeted forms of immune modulation, with the goal of uncoupling toxicity from the therapeutic effect of ICIs. Additional studies exploring the use of other biologic and targeted immunosuppressive treatment modalities for the management of cirAEs and irAEs more broadly are necessary.

Limitations of this study include its retrospective nature and limited sample size of the dupilumab-treated cohort. However, this is the largest study of dupilumab-treated cirAEs to date and the first to include robust comparator cohorts to investigate the impact of dupilumab on mortality in the ICI population. Future studies should confirm these findings among larger cohorts of ICI recipients and investigate the optimal time, dosing, and frequency for dupilumab therapeutic intervention in this population.

## Data Availability

All relevant data are available from the corresponding author: Yevgeniy R. Semenov. All summary data supporting the findings of this study are available within the article and/or its supplementary materials. The patient-level data generated for this study can only be shared per specific institutional review board requirements.

## Data Availability

The authors confirm that the data supporting the findings of this study are available within the article and/or its supplementary materials (available online).

## Declarations

YRS is an advisory board member or consultant and has received honoraria from Pfizer, Incyte Corporation, Sanofi, Galderma, Castle Biosciences, and Iovance Biotherapeutics. KLR is an advisory board member to SAGA Diagnostics, has received speaker fees from CMEOutfitters, MedScape, and BMS, and provides institutional support for the ATRIUM clinical trial. NRL is a consultant and has received honoraria from Bayer, Silverback, Fortress Biotech, and Synox Therapeutics outside the scope of the submitted work.

## Ethics approval

This study received approval from the Massachusetts General Brigham Institutional Review Board under protocol number 2020P002307.

## Competing interests

**N/A.**

## Funding

YRS is supported in part by the National Institute of Arthritis and Musculoskeletal and Skin Diseases of the National Institutes of Health (award number K23AR080791), the Department of Defense (award number W81XWH2110819), and the Melanoma Research Alliance Young Investigator Award. GW is supported by the National Cancer Institute of the National Institutes of Health (award number K99CA286966).

## Author’s Contributions

Study concept and design: GW, SK, NRL and YRS. Data collection: SK, GW, SX, CM, MT, EB, CL, and BL. Data analysis: GW, SK, YRS. Data interpretation: all authors. Drafting of the manuscript: GW, SK, NRL, YRS. Administrative, technical, or material support: YRS. Study supervision: NRL and YRS. GW, SK, NRL, and YRS directly accessed and verified the underlying data reported in the manuscript. All authors had access to the summary data reported in the study. This manuscript was written by the lead investigators and was reviewed and approved for publication by all co-authors.

## Acknowledgements

N/A

## List of Abbreviations

|  |  |
| --- | --- |
| **ICI** | Immune checkpoint inhibitors |
| **IrAE** | Immune related adverse event |
| **CirAE** | Cutaneous immune related adverse event |
| **CTLA-4** | Cytotoxic T-Lymphocyte Associated Protein 4 |
| **PD-1** | Programmed Death-1 |
| **PD-L1** | Programmed Death-Ligand 1 |
| **NCCN** | National Cancer Comprehensive Network |
| **IL-4** | Interleukin 4 |
| **IL-13** | Interleukin 13 |
| **FDA** | Food and Drug Administration |
| **IgE** | Immunoglobulin E |
| **CTCAE** | Common Terminology Criteria for Adverse Events |
| **CCS** | Charlson Comorbidity Score |
| **NOS** | Not otherwise specified |
| **CBC** | Complete blood count |
| **HR** | Hazard ratio |
| **CI** | Confidence interval |
| **Q1** | The first quartile |
| **Q3** | The third quartile |

## Notes

### Competing Interest Statement

The authors have declared no competing interest.

