## Supplemental Table 1 and 2 for "Long-term mortality outcomes among immunotherapy recipients treated with dupilumab for the management of cutaneous immune-related adverse events"

**Table S1.** Drugs trialed before initiation of dupilumab therapy.

| **Drug category** | **Drugs administered before initiation of dupilumab therapy for managing cutaneous immune-related adverse events** |
|---|---|
| Topical treatment | Betamethasone, clobetasol, triamcinolone, fluocinonide, dexamethasone swish and spit, magic mouth wash, nystatin rinse, lidocaine rinse, doxycycline swish and spit, mometasone, clobetasol solution, hydrocortisone, fluocinolone, econazole, calcipotriene, vitamin E, niacinamide, clindamycin gel, mupirocin, topical pimecrolimus, topical tacrolimus, silver sulfadiazine cream, Sarna, Aquaphor, CBD:THC Wonderbalm, camphor/menthol, Curel cream, CeraVe anti-itch cream, Eucerin, Vaseline |
| Hemorrhagic agents | Pentoxifylline |
| Oral vitamin | Nicotinamide |
| Opioid antagonist | Naltrexone |
| Oral systemic non-steroidal | Methotrexate, apremilast, mycophenolic acid |
| Anticonvulsants | Gabapentin, pregabalin |
| Oral antibiotics | Doxycycline, cefalexin, dapsone |
| Glucocorticoids | Prednisone, oral dexamethasone, intravenous methylprednisolone |
| Antihistamines | Diphenhydramine, cetirizine, hydroxyzine, fexofenadine, famotidine, loratadine |
| Biologics | Rituximab, omalizumab |
| Phototherapy | Narrow band ultraviolet B |
| Immunoglobulin | Intravenous immunoglobulin |
| Oral retinoid | Acetretin |

**Table S2.** Response to dupilumab stratified by morphology of cutaneous immune-related adverse events.

| | **Total (N=53)** | **Eczematous dermatitis (N=22)** | **Bullous pemphigoid (N=14)** | **Lichenoid dermatitis (N=7)** | **Morbilliform drug eruption (N=5)** |
|---|---|---|---|---|---|
| **Responder** | | | | | |
| Complete | 33 (62.3%) | 14 (63.6%) | 8 (57.1%) | 3 (42.9%) | 4 (80.0%) |
| Partial | 14 (26.4%) | 5 (22.7%) | 3 (21.4%) | 4 (57.1%) | 1 (20.0%) |
| No | 6 (11.3%) | 3 (13.6%) | 3 (21.4%) | 0 (0%) | 0 (0%) |
| | **Lichenoid dermatitis, Bullous pemphigoid (N=1)** | **Lichenoid dermatitis, eczematous dermatitis (N=1)** | **Lichenoid dermatitis, morbilliform drug eruption (N=1)** | **Radiation induced morphea ICI exacerbated (N=1)** | **Sclerodermoid reaction with morphea-profunda (N=1)** |
| **Responder** | | | | | |
| Complete | 1 (100%) | 1 (100%) | 1 (100%) | 0 (0%) | 1 (100%) |
| Partial | 0 (0%) | 0 (0%) | 0 (0%) | 1 (100%) | 0 (0%) |
| No | 0 (0%) | 0 (0%) | 0 (0%) | 0 (0%) | 0 (0%) |
